# Sex differences in placebo and antidepressant response to intranasal esketamine for treatment-resistant depression: a pooled participant analysis of randomized controlled trials

**DOI:** 10.1101/2025.06.26.25330356

**Authors:** Marie Huc, Sara Siddiqi, Mysa Myers, Ian Colman, Natalina Salmaso, Natalia Jaworska, Argel Aguilar-Valles

## Abstract

**Background:** Racemic ketamine and its enantiomer, esketamine, have emerged as fast-acting antidepressant options for individuals with treatment-resistant depression (TRD). Yet, despite growing clinical use, little is known about how sex assigned at birth shapes symptom-specific responses to these interventions, a critical gap in the move toward personalized psychiatry.

**Methods:** We conducted a pooled analysis of five randomized, double-blind, placebo-controlled trials in which adults with TRD received intranasal esketamine or placebo twice weekly for four weeks, alongside a newly initiated oral antidepressant. We evaluated the effects of sex assigned at birth on overall depression severity, measured via total Montgomery–Åsberg Depression Rating Scale (MADRS) scores, and across four symptom factors: sadness, negative thoughts, detachment, and neurovegetative symptoms. Rates of clinical response and remission were also analyzed by sex assigned at birth.

**Findings:** Overall, esketamine treatment improved total MADRS scores in both sexes; however, significant sex-specific patterns emerged. Females showed greater improvement in total MADRS scores than males towards the end of the trials, in both the placebo and esketamine arms. Females also showed more pronounced reductions in the sadness and detachment factors at the end of the trials, as well as in the neurovegetative factor on day 15, regardless of the treatment group. On the other hand, males showed a significant reduction in sadness symptoms after esketamine on day 2 of the treatment. Females had higher odds of responding, regardless of treatment arm, during later time points.

**Interpretation:** These findings reveal that sex assigned at birth influences overall antidepressant response and shapes the trajectory and symptom profile of improvement. Our findings emphasize the critical importance of incorporating sex assigned at birth as a key variable, essential for optimizing TRD treatment strategies and advancing individualized mental healthcare.

**Funding:** Canadian Institutes for Health Research.

## Introduction

Major Depressive Disorder (MDD) is one of the most prevalent mental health issues globally, with over 300 million estimated cases in 2021.^1,2^ MDD is a chronic and recurrent psychiatric disorder characterized by depressed mood, recurrent thoughts of death, feelings of worthlessness, social isolation and anhedonia, among other symptoms; it is also the leading risk factor for suicide.^3^ MDD presents significant challenges through its impact on quality of life, work productivity, healthcare systems and economic consequences.^1–3^ Thus, the discovery and evaluation of effective treatments is crucial, since existing first-line pharmacotherapies lead to remission in only about a third of patients within three months of treatment initiation.^4^ Indeed, many individuals fail to respond to more than two antidepressant interventions, rendering such patients as presenting with treatment-resistant depression (TRD).^5^ Recently, ketamine has emerged as a treatment option for patients with TRD. Ketamine (as a racemic mixture) and its two enantiomers, esketamine (S+) and arketamine (R-), are N-methyl-D-aspartate receptor (NMDAR) antagonists.^6^ Racemic ketamine was first introduced in medical practice as an effective intravenous anesthetic; however, at sub-anesthetic doses, it also induces a rapid reduction in depressive symptoms and suicidal ideation,^7^ within hours of a single dose.^6,8–11^ Nevertheless, repeated ketamine infusions are typically necessary for a cumulative and continued antidepressant response lasting up to 3 weeks,^9^ with over 80% of patients showing a response and remission following repetitive treatment.^12^ Despite this, ketamine’s use in treating TRD is limited by the high costs and the logistic complications linked to intravenous administration, combined with dissociative effects and abuse potential,^13^ raising some concerns about its widespread application and long-term safety in treating TRD.

The isolation of esketamine allowed for the development of lower doses administered intranasally, and was approved by the US Food and Drug Administration and in Europe in 2019, and by Health Canada in 2020 for TRD treatment, in combination with an oral antidepressant.^7,14,15^ Despite recent strides in the understanding of ketamine’s mechanism of action,^10,11,16–18^ it remains unclear whether sex assigned at birth (referred to as sex from here on) and gender modulate ketamine’s antidepressant response overall and on select symptoms of depressive disorders. This knowledge gap is particularly evident for esketamine, and addressing it is crucial to advancing the more optimized and targeted use of esketamine in the treatment of TRD.

Sex differences have been documented in the prevalence of MDD. Indeed, females are twice as likely to be affected by MDD compared to males.^8,9,19^ Furthermore, differences in symptom types, severity, and the presence and type of comorbidities have been observed between the sexes. For instance, women were often found to present more severe symptoms and are more likely to experience prolonged or recurrent depression at a younger age than men.^20^ Sex differences have also been noted in the antidepressant responses and side effects to medications such as selective serotonin reuptake inhibitors (SSRIs), serotonin-norepinephrine reuptake inhibitors (SNRIs) and tricyclic antidepressants (TCA).^8,9,14,19–21^ Concretely, some studies showed that men are more responsive to the TCA imipramine, while women respond better to SSRI antidepressants and, to a lesser extent, SNRIs. However, results are not always consistent.^20^

Research focusing on chronic recreational ketamine users demonstrated sex differences in withdrawal symptoms, with female patients experiencing more severe symptoms, including anxiety, dysphoria and cognitive impairments.^22^ Further, male participants demonstrate greater effects of ketamine on cardiac output and analgesia, while women exhibit faster drug clearance rates,^23^ likely due to sex differences in cytochrome P450 (CYP) enzyme expression and activity.^9^ These enzymes, which are regulated by estrogen and progesterone, play a role in ketamine’s metabolism, contributing to sex-specific differences in metabolic profiles and clinical outcomes, including varying levels of dissociative and psychotomimetic side effects.^9,24^ A recent review of preclinical literature found that female rodents have consistently more pronounced responses and sensitivity to ketamine (e.g., in dosage or magnitude of behavioural response).^9^ However, clinical data are variable, as another review reported that only 4 out of 27 reviewed articles showed evidence of sex differences in the antidepressant response to ketamine in TRD,^8^ while a previous meta-analysis showed no moderating effect of sex on intravenous racemic ketamine on depression symptoms.^25^

However, assessing sex differences has not been the primary objective of most existing studies on antidepressant use of ketamine or esketamine; further, individual studies tend to be underpowered to assess differences between males and females and/or have unbalanced samples.^8,9^ Thus, we conducted a focused analysis of sex differences in esketamine response in TRD, pooling data from multiple double-blinded, placebo-controlled clinical trials. We evaluated sex differences on the impact of esketamine treatment on specific depression symptom factors (i.e., sadness, negative thoughts, detachment and neurovegetative feelings). To our knowledge, this has never been assessed. Understanding such differences is crucial for optimizing treatment strategies and ensuring personalized, effective care and more equitable treatment across sexes.

## Methods

Data from clinical trials was accessed through the Yale Open Data Access (YODA) Project after approval by their data-sharing committee. While the initial proposal to YODA (see Supplementary file 1) outlined a meta-analysis focused on ketamine in general, the approach was modified to conduct a pooled analysis specifically focusing on esketamine. Most trials deposited in the YODA database were on esketamine, enabling greater homogeneity and a more suitable focus. In addition, our analysis plan modification was driven by the necessity to combine and analyze the results of the trials while maintaining adequate sample sizes at each visit time. Each of the studies analyzed was previously approved by their respective Research Ethics Boards (REB), and written consent was obtained from all participants. The studies are registered on clinicaltrials.gov; the YODA program allowed us to access eleven unique trials: 54135419SUI3001 (or ASPIRE-1, Trial ID: NCT03039192),^26^ ESKETINTRD3004 (or SUSTAIN-2, Trial ID: NCT02497287),^27^ ESKETINTRD3002 (or TRANSFORM-2, Trial ID: NCT02418585),^28^ ESKETINTRD3006 (Trial ID: NCT03434041),^29^ 54135419TRD2005 (Trial ID: NCT02918318),^30^ ESKETINTRD3001 (or TRANSFORM-1, Trial ID: NCT02417064),^31^ ESKETINTRD3005 (or TRANSFORM-3, Trial ID: NCT02422186),^32^ ESKETINTRD2003 (or SYNAPSE, Trial ID: NCT01998958),^33^ ESKETINTRD3003 (or SUSTAIN-1, Trial ID: NCT02493868),^34^ 54135419SUI3002 (or ASPIRE-II, Trial ID: NCT03097133),^26^ ESKETINSUI2001 (Trial ID: NCT02133001),^35^ ESKETIVTRD2001 (Trial ID: NCT01640080).^36^ This pooled analysis study was approved by Carleton University’s REB (#119295).

The full protocol of each of these trials was reviewed by two independent reviewers, who assessed them according to our inclusion criteria, and conflicts were resolved by consensus. Inclusion criteria were: a double-blind, randomized controlled trial, the population of interest (aged 18+), TRD with esketamine treatment intervention being compared to placebo/antidepressant treatment, and the outcome of interest/data available for assessment of ketamine’s efficacy on measures of depression using a standardized questionnaire. One trial was excluded from analysis because it did not have the targeted study design (open-label [ESKETINTRD3004]), one trial assessed a different outcome (continuation/discontinuation of treatment on relapse [ESKETINTRD3003]), and one trial had a different route of esketamine administration (intravenous), along with differences in treatment timeline; as such, it was excluded to reduce heterogeneity (ESKETIVTRD2001). Exploratory analysis was conducted for the remaining nine studies; however, four of the trials were excluded from analysis due to unavailable outcome data (54135419SUI3001, 54135419SUI3002, ESKETINSUI2001, ESKETINTRD2003). As such, a total of five studies were included in the final pooled analysis to assess the role of sex differences on the antidepressant effects of esketamine for the treatment of TRD (ESKETINTRD3002, ESKETINTRD3006, 54135419TRD2005, ESKETINTRD3001, ESKETINTRD3005). See the supplementary methods for complete details regarding data extraction from each trial, which was conducted in May 2024. Further information on these trials is reported elsewhere.^37–41^

Briefly, the final five trials administered esketamine to patients with TRD through a nasal spray in addition to a newly initiated antidepressant. All trials were randomized, double-blind, placebo-controlled (details below), multicenter studies. The studies were conducted between 2015 and 2021. Participants aged 18 to 64 received a diagnosis following the Diagnostic and Statistical Manual of Mental Disorders – Fifth Edition (DSM-5)^42^ for single-episode or recurrent moderate to severe MDD, without psychotic features, based on clinical assessment and confirmed by the Mini International Neuropsychiatric Inventory.^43^ At the start of the trials, participants were diagnosed with TRD, defined as no clinically meaningful improvement after at least two different antidepressant agents were used as prescribed. Main exclusion criteria (with some variations within each trial) included recent deep brain stimulation or vagal nerve stimulation during the current depressive episode, current diagnoses of psychotic disorders, bipolar disorders or related disorders, or co-occurring substance or alcohol use disorders. Participants in the trials were randomly assigned to doses of 28mg, 56mg or 84mg esketamine or a placebo dose of 0·001mg/mL denatonium benzoate bittering agent.

For trials ESKETINTRD3002, ESKETINTRD3006, ESKETINTRD3001 and ESKETINTRD3005, participants in the experimental arm self-administered esketamine intranasally twice per week for 4 weeks as a flexible dose regimen in the Double-Blind Induction Phase. Doses were 28, 56 or 84 mg. In addition, participants simultaneously initiated a new, open-label oral antidepressant (i.e., duloxetine, escitalopram, sertraline, or venlafaxine extended-release [XR]) on Day 1, which was continued throughout the Double-Blind Induction Phase (DBIP). Trial 54135419TRD2005 also dosed patients in the experimental arm twice a week for 4 weeks, but the doses were fixed (28, 56, or 84 mg) for the duration of the DBIP. All included trials monitored symptoms regularly using standardized and clinician-rated scales (details below).

### Outcome measures

The primary outcome of interest in this study was the change in Montgomery-Asberg Depression Rating Scale (MADRS)^44^ scores between the sexes from screening to each subsequent return visit following esketamine administration (at 1, 8, 15, 22, and 28 days into treatment). Secondary outcomes included the assessment of the change in MADRS score from baseline to each return visit between the sexes for each component in the Four Factor model (i.e., sadness, negative thoughts, detachment, and neurovegetative [details below] to more granularly assess putative sex-specific features);^45^ the proportion of responders/non-responders (details below) per sex, and the proportion of patients in remission at each post-treatment visit per sex.

The factor “sadness” was assessed through summing scores for items 1 and 2 of the MADRS, corresponding to *Apparent Sadness*, which reflects observable signs of gloom and despair in speech, facial expression, and posture, and *Reported Sadness*, which captures an individual’s verbal account of feelings of hopelessness.^45,46^ The “neurovegetative” factor was measured through a sum of the MADRS item 3, *Inner Tension*, which reflects feelings of discomfort and escalating anxiety; item 4, *Reduced Sleep*, representing a decrease in sleep duration, and item 5, *Reduced Appetite*, which captures a loss of desire for food or the need to force oneself to eat.^45,46^ “Detachment” was assessed using a sum of item 6, *Concentration Difficulties*, reflecting challenges in focusing or organizing thoughts; item 7, *Lassitude*, representing difficulty in initiating or performing daily activities, and item 8, *Inability to Feel*, capturing a reduced emotional response or diminished interest in typically enjoyable.^45,46^ Finally, the factor “negative thoughts” was defined as the sum of item 9, *Pessimistic Thoughts*, reflecting feelings of guilt or inferiority, and *Suicidal Thoughts*, which encompass thoughts of death, and suicidal ideation or preparations.^45,46^ Responders were classified according to an improvement from baseline greater than 50% on the MADRS scale (non-responder: less than/including 49% score decrease); remission was defined by a MADRS score <10.^47^

### Statistical Analyses

Data from all five trials were pooled, and descriptive statistics (mean, standard deviation, counts, and percentages) were used to summarize demographic variables for participants at the baseline/screening visit, prior to receiving esketamine and placebo. A two-way ANOVA was conducted for each visit time, comparing the mean change in MADRS scores (from baseline) across sex, treatment (ketamine vs. placebo), and the interaction between sex and treatment. The average change in MADRS score was defined by the mean difference between the total score per visit and baseline score within each group. Next, a similar two-way ANOVA was conducted to assess the mean change in score for each level of the Four Factor Model.^45^ Ranked values were used for the ANOVAs when assumptions were violated. Partial effect sizes were determined and reported.

A logistic regression model was fitted to compare responders and non-responders as outcome measures, with treatment arm and sex as predictors at each return visit following baseline. A deviance test was conducted to assess the role of sex-treatment arm interaction; if non-significant, then the main model was selected. Logistic regression with a deviance test was repeated to model remission versus non-remission. Significance was considered when p<0·05. All analysis was conducted in RStudio (R version 4·3·0).

## Results

### Sex-specific effects of placebo and esketamine on the change in depressive symptoms

Participant characteristics (440 Placebo; 576 Esketamine, across n=5 studies) are presented in Table 1 and for each visit in Supplementary Table 1. We first investigated whether esketamine had sex-specific effects on the change in the total MADRS score from baseline to 1, 2, 8, 15, 22 and 28 days into the DBIP (Fig. 1). A main effect of the treatment arm (placebo vs. ketamine) was evident from the second visit and onwards (Fig. 1; day 2: F_(1,866)_=16·81, *p<*0·001, partial η²<0·001; day 8: F_(1,964)_=14·18, *p<*0·001, partial η²=0·002; day 15: F_(1,944)_=17·56, *p<*0·001, partial η²=0·004; day 22: F_(1,920)_=21·69, *p<*0·001, partial η²=0·005; and day 28: F_(1,907)_=26·03, *p<*0·001, partial η²=0·007), but not on the first day after treatment onset (Fig. 1; day 1: F_(1,226)_=0·03, *p*=0·87, partial η²<0·001).

**Figure 1.**
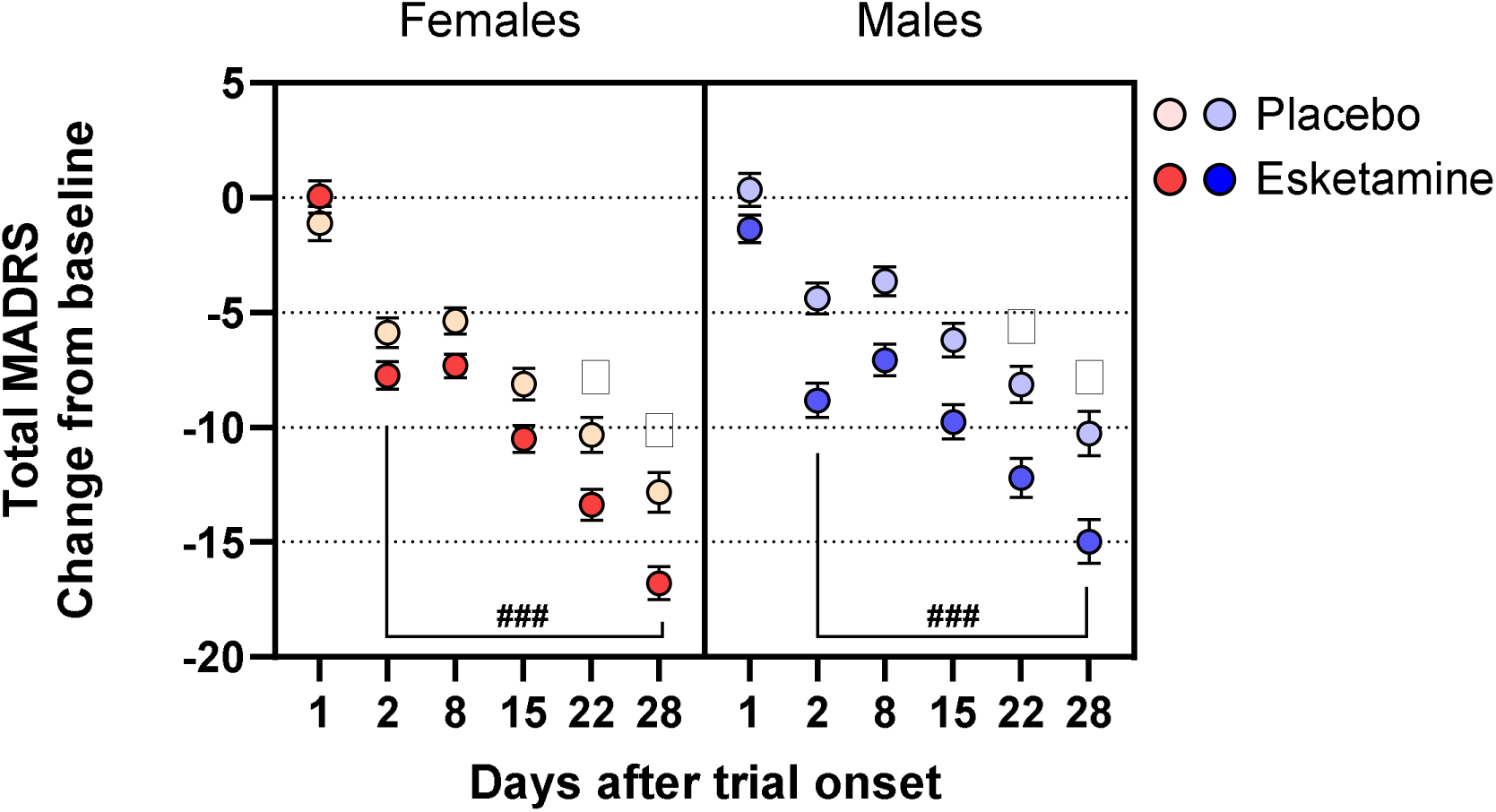
Total change in MADRS score across study time. (Right) Changes in MADRS scores for females from baseline to days 1-28 into the double-blind induction phase (DBIP) of the trials and across participants in the placebo or esketamine groups. (Left) Changes in MADRS scores for males from baseline to days 1-28 in the DBIP across participants in the placebo or esketamine groups ### p<0·001, Main effect of treatment arm at the indicated time points; ⋇ p<0·05, Main effect of sex at the indicated time points.

**Table 1.** Participant Characteristics (means and standard deviation presented)

|  | **Placebo**<br>**Group**<br>**(n=440)** | **Esketamine**<br>**Treatment Group**<br>**(n=576)** |
| --- | --- | --- |
| **Age\* (year)** | 36.6±12.0 | 36.6±11.6 |
| **BMI\*\*** | 27.5±6.1 | 27.4±5.9 |
| **Females (%)** | 251 (57.0%) | 353 (61.2%) |
| **Ethnicity (%)** |  |  |
| Hispanic or Latino | 43 (9.8%) | 73 (12.7%) |
| Not Hispanic or Latino | 256 (58.2%) | 358 (62.2%) |
| Not Reported or Unknown | 141 (32.0%) | 145 (25.2%) |
| **Baseline MADRS Score** | 36.4±4.8 | 36.6±5.0 |

BMI: Body mass index
\* n=147 and 161 participants with data in the placebo and esketamine groups, respectively
\*\* n=419 and 540 participants with data in the placebo and esketamine groups, respectively

A main effect of sex was found on days 22 (Fig. 1; F_(1,920)_=4·75, *p=*0·03, partial η²=0·02) and 28 days after treatment onset (Fig. 1; F_(1,907)_=6·08, *p=*0·01, partial η²=0·03), in which females (F) had greater decreases in the MADRS score than males (M), regardless of treatment arm (day 22: F=-12·07 ± 0·51, M=-10·36 ± 0·60; day 28: F=-15·12 ± 0·55, M=-12·80 ± 0·68). For all other days, the effect of sex was not significant (Fig. 1; *p*>0·05). Finally, a significant interaction between treatment arm and sex was found at the first visit after treatment onset (F_(1,226)_=4·11, *p*=0·04, partial η²=0·02).

### Sex specific effects on depressive symptoms factors in esketamine trials

*Sadness factor.* Results per factor are summarized in Fig. 2. Similar to the total MADRS scores (Fig. 1), a significant effect of the treatment arm on changes in MADRS sadness factor was observed for days 2, 8, 15, 22 and 28 after DBIP onset (Fig. 2A), reflecting esketamine’s effects in reducing sadness (day 2: F_(1,866)_=22·93, *p<*0·001, partial η²=0·001; day 8: F_(1,964)_=12·95, *p<*0·001, partial η²=0·003; day 15: F_(1,944)_=11·28, *p<*0·001, partial η²=0·004; day 22: F_(1,920)_=18·20, *p<*0·001, partial η²=0·007; and day 28: F_(1,907)_=26·22, *p<*0·001, partial η²=0·007). The effect of the treatment arm was not significant on the first day after treatment onset (F_(1,226)_=0·36, *p*=0·55, partial η²<0·001). Similar to the total MADRS score, there was a main effect of sex on days 22 and 28 for the MADRS sadness factor (day 22: F_(1,920)_=6·18, *p=*0·01, partial η²=0·02; day 28 F_(1,907)_=5·9468, *p=*0·02, partial η²=0·03), with greater MADRS sadness score reductions in females compared to males, in both treatment arms (day 22: F=-3·19 ± 0·14, M= -2·66 ± 0·17; day 28: F=-3·94 ± 0·16, M=-3·26 ± 0·19). There was a significant treatment × sex interaction on day 2 (F_(1,866)_=5·15, *p=*0·02, partial η²=0·006), but not for any other time point (*p*>0·05). At this time point, there was a distinction between males treated with esketamine vs. placebo (Fig. 2A, right panel); this effect was absent in females.

**Figure 2.**
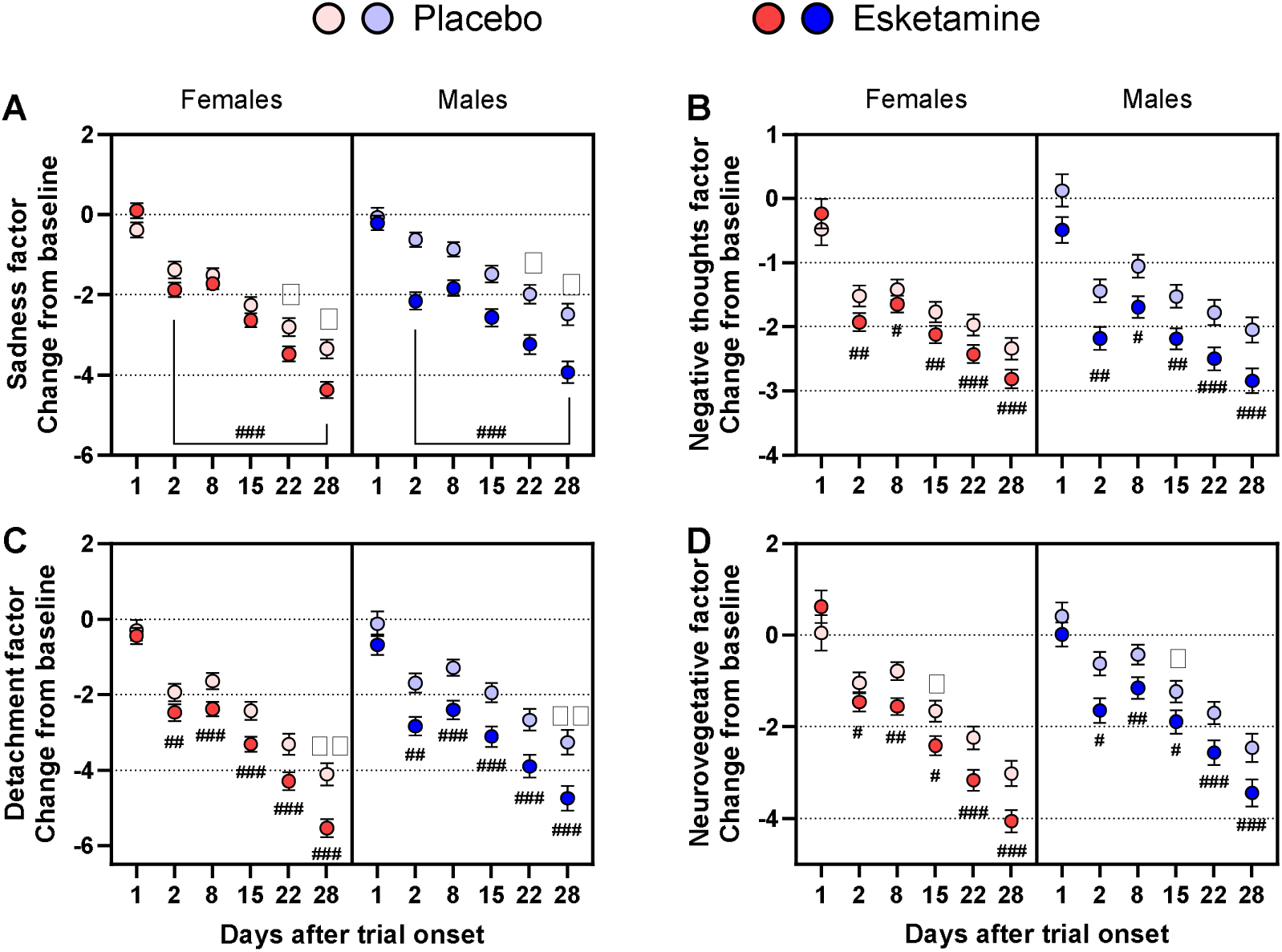
Effects of sex and treatment on MADRS factors. Panels represent results across each MADRS factor: (A) Sadness, (B) Negative Thoughts, (C) Detachment, (D) Neurovegetative. (Right of each panel) Changes in MADRS scores for females from baseline to day 1-28 across participants in the placebo and esketamine groups. (Left of each panel) Changes in MADRS scores for males from baseline to day 1-28 across participants in the placebo to esketamine groups. # p<0·5; ## p<0·01; ### p<0·001, Main effect of treatment arm at the indicated time points. ⋇ p<0·05; ⋇⋇ p<0·01, Main effect of sex at the indicated time points.

**Figure 3.**
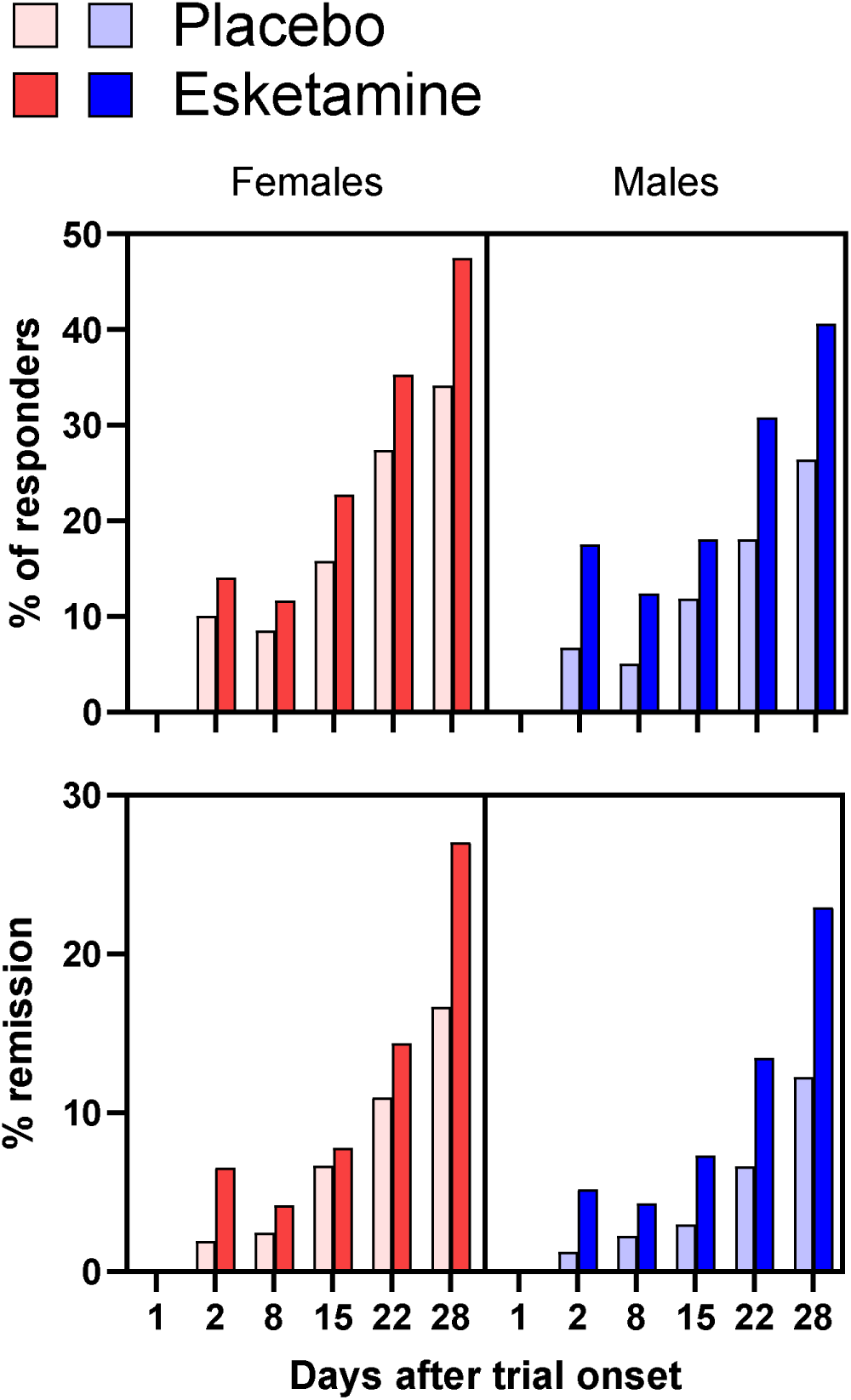
Rates of response and remission to esketamine. (A) The percentage of responders at each time point, across treatment arms and sex. (B) The percentage of participants in remission at each time point, across treatment arms and sex. Males are indicated in blue, and females in red.

*Negative thoughts factor.* For the negative thoughts factor, the main effect of treatment was significant for days 2 through 28 (Fig. 2B; day 2: F_(1,866)_=9·63, *p=*0·002, partial η²<0·001; day 8: F_(1,964)_=5·90, *p=*0*·*02, partial η²<0·001], day 15: F_(1,944)_=6·79, *p=*0·009, partial η²<0·001; day 22: F_(1,920)_=10·99, *p<*0·001, partial η²<0·001; day 28: F_(1,907)_=13·41, *p<*0·001, partial η²<0·001), but not for 1 day after treatment (F_(1,226)_=0·33, *p*=0·57, partial η²=0·002). There was no effect of sex or any significant interactions for the negative thoughts factor.

*Detachment factor.* There was a main effect of the treatment arm on MADRS detachment factor changes on days 2 through 28 (Fig. 2C; day 2: F_(1,866)_=8·41, *p=*0·004, partial η²<0·001; day 8: F_(1,964)_=16·11, *p<*0*·*001, partial η²<0·001; day 15: F_(1,944)_=15·90, *p<*0·001, partial η² =0·002; day 22: F_(1,920)_=16·43, *p<*0·001, partial η²=0·004; day 28: F_(1,907)_=25·89, *p<*0·001, partial η²=0·009), but not 1 day after treatment onset (F_(1,226)_=1·34, *p*=0·25, partial η²<0·001). Furthermore, 28 days after treatment onset, a main effect of sex on changes in the detachment factor was noted (Fig. 2C; F_(1,907)_=7·81, *p=*0·005, partial η²=0·03], with a greater reduction in the MADRS detachment factor in females (F=-4·94 ± 0·19, M=-4·07 ± 0·23).

*Neurovegetative factor.* Similar to sadness and detachment factors, apart from day 1 (F_(1,226)_=0·00, *p*=1·0, partial η²<0·001), there was a main effect of treatment on MADRS neurovegetative factor changes at all time points (Fig. 2D; day 2: F_(1,866)_=6·14, *p=*0·01, partial η²<0·001), day 8: F_(1,964)_=9·95, *p=*0*·*002, partial η²=0·003; day 15 F_(1,944)_=6·24, *p=*0·013<0·001, partial η²=0·004], day 22: F_(1,920)_=152·79, *p<*0·001, partial η²=0·003; day 28: F_(1,907)_=13·13, *p<*0·001, partial η²=0·003). There was also a main effect of sex on neurovegetative factor changes only 15 days after treatment onset (Fig. 2D; F_(1,944)_=3·99, *p=*0·046, partial η²=0·007); this was attributed to a larger reduction in females than males, regardless of treatment arm (F=-2·10 ± 0·16, M=-1·59 ± 0·18).

### Sex specific effects on treatment on response and remission in the esketamine trials

The logistic regression analysis revealed that response was significantly associated with treatment arm (placebo or esketamine) at days 2 (Beta Estimate=0·66, OR (95% CI)= 1·93 (1·25, 3·00), p<0·001), 8 (Beta Estimate=0·57, OR (95% CI)=1·77 (1·12, 2·78), p=0·014), 15 (Beta Estimate=0·46, OR(95% CI) =1·59 (1·12, 2·25), p=0·009), 22 (Beta Estimate=0·49, OR (95% CI) =1·63 (1·21, 2·19), p=0·001] and 28 post-treatment (Beta Estimate=0·59, OR (95% CI)=1·80 (1·37, 2·37), p<0·001). Further, on days 22 and 28, females had a higher odds of treatment response compared with males (p<0·05). However, no sex-treatment interactions were found.

There were also significant regressions on the impact of treatment on remission at days 2 (Beta Estimate=1·34, OR (95% CI) =3·84 (1·58, 9·33), p=0·003), 22 (Beta Estimate=0·47, OR(95% CI) = 1·60 (1·05, 2·43), p=0·03) and 28 post-treatment (Beta Estimate=0·66, OR (95% CI) =1·94 (1·38, 2·73), p=0·0001). There were no sex differences or sex-treatment interactions at any time point for remission.

## Discussion

This study investigated sex differences in response to esketamine treatment for TRD. As expected, we found greater improvements in the esketamine compared to the placebo arm on total MADRS scores and across symptom factor scores (i.e., sadness, negative thoughts, detachment and neurovegetative factors). Similarly, those in the ketamine arm had greater rates of remission and response, which is consistent with existing literature showing robust antidepressant effects within hours of initiating ketamine, and lasting for several weeks with repeated exposure.^8,9^ Critically, females showed greater total MADRS improvement than males at days 22 and 28 following treatment onset, regardless of treatment arm.

Given this finding, we employed a symptom-based approach, which enables a more granular assessment of putative sex differences. Females showed more pronounced improvements in the sadness factor on days 22 and 28, in the detachment factor on day 28, and in the neurovegetative factor on day 15, also regardless of treatment. Males, on the other hand, showed a significantly greater reduction in sadness after esketamine vs. placebo on day 2; this effect was absent among females. Women show greater MDD symptom severity than men,^20^ and sex differences in symptoms in chronic ketamine users have been observed.^22^ As such, our analyses focusing on specific symptoms reveal a pattern of response whereby males respond more strongly to esketamine acutely, but over time, females exhibit a stronger response to both placebo and esketamine. Sex differences were also observed on days 22 and 28 in responders, with a greater odds of females responding in both the placebo and ketamine arms of the trials, relative to males.

While sex differences in the antidepressant effect of ketamine have been shown in some studies, typically with a higher response in females, others show no sex differences.^8,14,25^ Of note, most individual studies have small sample sizes, resulting in an underpowered analysis of sex differences.^8^ To our knowledge, only one study focused on assessing differences between males and females in a human sample^14^ and showed no significant sex differences in esketamine response, although a smaller sample size compared to our pooled analysis may have masked sex differences.

Our analyses showed greater improvements in females than in males, in both the placebo and esketamine arms, for sadness on days 22 and 28, detachment on day 28, and neurovegetative factors on day 15. Previous work indicates that women might be more sensitive to negative emotions, particularly sadness,^48–50^ and show differential brain activity in emotion processing.^51,52^ It is thus possible that increased emotional sensitivity could make females more responsive to therapeutic interventions. In addition, previous studies have shown a wider range of maladaptive responses (i.e., maintaining or increasing sadness) in females compared to males, as well as increased rumination, avoidance, and wishful thinking.^53–55^ In contrast, males seem to use substance use and suppression of emotional expression to cope with sadness.^53–55^ Further, symptoms such as changes in appetite, sleep, fatigue, somatic anxiety and psychomotor disturbance are usually over-represented in females with MDD compared to males.^56^ Therefore, women may experience greater symptoms of sadness, detachment and neurovegetative factors, in general, but also more pronounced responses to improvement of these symptoms. While our results showed that these improvements were more pronounced in women, they may partly reflect higher baseline severity; however, our analyses were based on changes from baseline scores (i.e., mean changes between baseline and treatment timepoints) to account for potential confounds related to baseline severity. Importantly, these greater responses were observed in the placebo and esketamine treatment arms (although esketamine arm had greater responses at several time points, regardless of sex), indicating a greater sensitivity to treatment in general, as all these patients in either arm had SSRI or SNRIs co-administered. While results are inconsistent across populations, previous research observed that women tend to respond better to SSRI antidepressants and, to a lesser extent, SNRIs.^20^

The analysis of effects on the sadness MADRS factor also revealed that females showed generalized score reduction across both treatment arms. In contrast, males’ responses for the sadness factor were more treatment-specific, potentially reflecting differences in how males and females respond to treatment.

We did not observe any sex differences in the decreases in the negative thoughts factor (i.e., pessimistic and suicidal thoughts), as esketamine decreased it equally in both sexes. Previous research suggested that although females attempt suicide more often, males have higher suicide mortality, but negative/suicidal thoughts are a risk factor common to both sexes.^57^ In this regard, our analysis indicated that esketamine-induced improvement in the negative thoughts factor is not influenced by biological or psychosocial factors related to sex.

One of our key observations is the greater response in the female placebo group, compared to the placebo-treated males. While some reviews and meta-analyses observed higher placebo response in females in treatments for restless leg syndrome,^58^ schizophrenia,^59^ and bipolar disorder.^60^ This effect is inconsistent across antidepressant treatments,^61^ and other pharmacotherapies.^62^ Notably, strong placebo effects may obscure the effectiveness and/or response to treatment, as other factors, such as expectation, can also impact treatment response. Interestingly, placebo effects rely on brain regions involved in affective processes, such as emotion regulation and fear management, including the amygdala, orbitofrontal cortex, and dorsal cingulate cortex;^63^ sex differences have been observed in the brain activity of these areas in individuals with MDD.^50,64^ Therefore, sex-related differential activity in these regions may contribute to a stronger placebo response in females. Notably, participants in the placebo arms of the pooled clinical trials were initiating a new SSRI or SNRI at trial onset; therefore, it is possible that the greater response in this group among females was linked to their response to the other antidepressant agent.

### Limitations and future directions

This pooled analysis, including a large sample size, assessed sex differences in specific symptom factors of MDD and demonstrated the importance of monitoring sex-specific responses at different time points across treatments. Nevertheless, our findings should be interpreted cautiously due to some heterogeneity in the trials, such as the variability in dosing regimens. Furthermore, other factors, such as side effects, should also be considered, as they may influence the results. For instance, while men report experiencing more cognitive and psychotic-related symptoms, women tend to report more nausea and headaches following ketamine and esketamine treatments.^9,21^ Importantly, our analyses are limited by the lack of collection of gender-related variables. Additionally, the sample exhibited heterogeneity in terms of age and ethnicity, though this does increase generalizability. Limited power and a lack of data precluded our ability to control for these important covariates, as has been observed with neurodegenerative disorders.^65^ Further studies should aim to explore sex differences, considering adverse effects and demographic factors (e.g., ethnicity, age, gender). Overall, our findings demonstrate the need to incorporate sex as a variable in treatment strategies and clinical trials for depression, with particular attention to placebo effects and symptom-specific factors to tailor treatments to individuals (e.g., by adjusting monitoring dose and frequency of treatment based on sex, considering that females seem to respond better).

## Author contributions

Authors participated in this research by contributing to its conceptualization (AAV, MH, MM, NJ, SS), methodology (AAV, MH, MM, NJ, NS, IC, SS), statistical analyses (AAV, MH, SS, IC), draft preparation (AAV, MH, SS) and reviewing and editing (AAV, MH, SS, NJ, NS, MM, IC). All authors approved the submitted version.

## Declaration of Interest

AAV is a scientific advisor for BetterLife Pharma and previously held a research contract with Gilgamesh Pharma. None of these companies was involved in the current study.

## Data Availability

All data produced in the present work are contained in the manuscript

## Acknowledgements

This work was funded by a CIHR (Canadian Institutes of Health Research) project grant to AAV. The funding agency was involved in the execution of the current study.

This study, carried out under YODA Project #2023-5149, used data obtained from the Yale University Open Data Access Project, which has an agreement with JANSSEN RESEARCH & DEVELOPMENT, L.L.C. The interpretation and reporting of research using this data are solely the responsibility of the authors and do not necessarily represent the official views of the Yale University Open Data Access Project or JANSSEN RESEARCH & DEVELOPMENT, L.L.C.

## Notes

### Clinical Trial

NCT02418585; NCT03434041; NCT02417064; NCT02422186; NCT03097133

### Author Declarations

Data from clinical trials was accessed through the Yale Open Data Access (YODA) Project after approval by their data-sharing committee. Each of the studies analyzed was previously approved by their respective Research Ethics Boards (REB), and written consent was obtained from all participants. This pooled analysis study was approved by Carleton University's REB (#119295).

