## Supplementary methods for "Sex differences in placebo and antidepressant response to intranasal esketamine for treatment-resistant depression: a pooled participant analysis of randomized controlled trials"

*Data Extraction from trials in YODA*

To prepare the raw data from each trial for the pooled analysis, variables of interest were extracted, and a data frame was created for each study on May 6, 2024. All data extraction was conducted in RStudio (R version 4.3.0 [2023-04-21 ucrt]), the haven,^1^ dplyr,^2^ and tidyverse^3^ packages were used to read/import and organize the data, respectively. Firstly, demographic variables (*Sex, Age, Ethnicity and Experimental Arm*) were selected; all screen failures were removed from the analysis. Data values presented in Table 1 for Age, Ethnicity, and BMI are based on available data; missing data were excluded from these summary estimates. Next, data pertaining to total MADRS score and study visit date were extracted for participants during the experimental phase of the trial. Furthermore, MADRS scores corresponding to the Four Factor Model (specific questions on the MADRS scale) were calculated for participants in the experimental phase of the study. Participants with multiple scores for a specific question on the MADRS scale at a given date, or those with early withdrawal, were removed from the analysis. This process was repeated to determine all relevant scores for participants during the study's screening phase. Values for screening considered were from baseline, determined by sorting the earliest date of records (participants with multiple screening measures recorded on the same earliest date were removed from analysis). Data for screening and experimental phases were combined by matching Subject ID. Next, we determined the change in total MADRS score from baseline at each Visit Date, and a complete case analysis for change in scores was applied. Next, the change in each level of the Four Factor Model from screening at each visit date was determined. In addition, BMI data collected were extracted; n=1 study was missing all BMI data, and the mean presented in Table 1 is based on the available data. Finally, demographic data, MADRS scores/change in scores and BMI data were combined; thereby creating a data frame for each trial. Data frames of each included trial were pooled to conduct further analyses (2005 n=285, 3001 n=1626, 3002 n=1148, 3005 n=534 , 3006 = 1272), any remining duplicates of visit date-participant combinations were removed (n=2), any unscheduled visits were not considered (n=1), participants under 18 were excluded (n=10), the final pooled set had n=4852 unique participant-visit records (Supplementary Table 1).

*Data Analysis*

A non-parametric (rank transformed) linear mixed effects model was used to test for the significance of a three-way interaction of visit, treatment arm and sex on the outcome of final visit MADRS score with a hierarchically well formulated model and adjusting for screening/baseline MADRS, and including a random participant intercept; continuous predictors were centered, and the screening MADRS was rank transformed to retain consistency with outcome transformation. This model was developed with the lmerTest package,^4^ and assumptions of independence of observations within groups of treatment arm, constant variance, no influential outliers (broom.mixed package^5^) and no multicollinearity (car package^6^) was verified. Influential outliers prior to conducting the logistic regression analysis were assessed with the broom package.^5^ Assumptions of independent observations and multicollinearity were also verified for logistic regression. In addition, assumptions of normality and constant variance were tested prior to conducting ANOVAs; rank transformations were applied to the outcome when violations were present. Adjustment for multiple comparisons was not conducted, as participant repeated measures are not independent.

**Supplementary Table 1.** Participant Characteristics (means and standard deviation presented)^a^

| Variables | **Placebo Arm** | | | | | | **Esketamine Treatment Arm** | | | | | |
| --- | --- | --- | --- | --- | --- | --- | --- | --- | --- | --- | --- | --- |
|  | Visit 1  (n=109) | Visit 2  (n=371) | Visit 8  (n=423) | Visit 15  (n=409) | Visit 22  (n=403) | Visit 28  (n=397) | Visit 1  (n=121) | Visit 2  (n=499) | Visit 8  (n=545) | Visit 15  (n=539) | Visit 22  (n=521) | Visit 28  (n=514) |
| Age (SD)* | 36·9 (12·3) | 36·6 (12·0) | 36·5 (11·9) | 36·7 (11·9) | 36·9 (11·9) | 37·2 (12·0) | 37·0 (12·3) | 36·7 (11·6) | 36·8 (11·6) | 36·7 (11·6) | 36·5 (11·6) | 36·6 (11·7) |
| BMI (SD)* | 26·2 (5·8) | 27·1 (5·9) | 27·5 (6·1) | 27·6 (6·0) | 27·6 (6·0) | 27·6 (6·1) | 26·3 (6·2) | 27·2 (6·0) | 27·4 (6·0) | 27·5 (6·0) | 27·6 (6·0) | 27·6 (6·1) |
| Female (%) | 60 55·0% | 208 (56·1% | 245 57·9% | 240 58·7% | 237 58·8% | 234 58·9% | 68 56·2% | 305 61·1% | 335 61·5% | 334 62·0% | 320 61·4% | 322 62·6% |
| Ethnicity (%) |  |  |  |  |  |  |  |  |  |  |  |  |
| Hispanic or Latino | 3  2·8% | 38 10·2% | 41  9·7% | 41 10·0% | 39  9·7% | 40 10·1% | 4  3·3% | 64 12·8% | 71 13·0% | 70 13·0% | 68 13·1% | 68 13·2% |
| Not Hispanic or Latino | 45 41·3% | 194 52·3% | 251 59·3% | 244 59·7% | 241 59·8% | 238 59·9% | 44 36·4% | 296 59·3% | 338 62·0% | 333 61·8% | 323 62·0% | 321 62·5% |
| Not Reported/ Unknown | 61  56·0% | 139 37·4% | 131 31·0% | 124 30·3% | 123  30·5% | 119  30·0% | 73  60·3% | 139 27·9% | 136 25·0% | 136 25·2% | 130 25·0% | 125 24·3% |
| Baseline MADRS Score | 36·4 (5·0) | 36·7 (4·8) | 36·4 (4·9) | 36·4 (4·9) | 36·5 (4·9) | 36·5 (4·9) | 37·3 (5·6) | 36·6 (5·0) | 36·6 (5·0) | 36·6 (5·1) | 36·6 (5·0) | 36·5 (5·0) |

^a^ One participant with data at visit 4 included in sample, demographic data not shown

*Data not available for all participants, missing data excluded from summary (not from main analysis)
