## Supplementary file 1 for "Sex differences in placebo and antidepressant response to intranasal esketamine for treatment-resistant depression: a pooled participant analysis of randomized controlled trials"

### Principal Investigator

**First Name:** Mysa
**Last Name:** Myers
**Degree:** Bachelor of Cognitive Science Honours
**Primary Affiliation:** Carleton University
**Phone number:**
**Address:**

**City:** Ottawa
**State or Province:** Ontario
**Zip or Postal Code:** K1S 5B6
**Country:** Canada

### General Information

**Key Personnel (in addition to PI):**
**First Name:** Mysa
**Last name:** Myers
**Degree:** MSc (student), Neuroscience
**Primary Affiliation:** Carleton University
**SCOPUS ID:** 57760227400

**First Name:** Marie
**Last name:** Huc
**Degree:** PhD (candidate), Neuroscience
**Primary Affiliation:** Carleton University
**SCOPUS ID:**

**First Name:** Argel
**Last name:** Aguilar-Valles
**Degree:** PhD, Neuroscience
**Primary Affiliation:** Carleton University
**SCOPUS ID:** 56013671900

**Are external grants or funds being used to support this research?:** No external grants or funds are being used to support this research.
**How did you learn about the YODA Project?:** Colleague

### Conflict of Interest

https://yoda.yale.edu/system/files/sv_6m4tghhxg7w7uxe-r_1dhzjmrcjbhhheb.pdf
https://yoda.yale.edu/system/files/sv_6m4tghhxg7w7uxe-r_3ja7ccfacahcpze.pdf
https://yoda.yale.edu/system/files/sv_6m4tghhxg7w7uxe-r_2xppmbw7vwuw8pq1.pdf

### Certification

**Certification:** All information is complete; I (PI) am responsible for the research; data will not be used to support litigious/commercial aims.
**Data Use Agreement Training:** As the Principal Investigator of this study, I certify that I have completed the YODA Project Data Use Agreement Training

1. NCT02417064 - ESKETINTRD3001 - A Randomized, Double-blind, Multicenter, Active-controlled Study to Evaluate the Efficacy, Safety, and Tolerability of Fixed Doses of Intranasal Esketamine Plus an Oral Antidepressant in Adult Subjects With Treatment-resistant Depression
2. NCT02418585 - ESKETINTRD3002 - A Randomized, Double-blind, Multicenter, Active-controlled Study to Evaluate the Efficacy, Safety, and Tolerability of Flexible Doses of Intranasal Esketamine Plus an Oral Antidepressant in Adult Subjects With Treatment-resistant Depression
3. NCT02422186 - ESKETINTRD3005 - A Randomized, Double-blind, Multicenter, Active-controlled Study to Evaluate the Efficacy, Safety, and Tolerability of Intranasal Esketamine Plus an Oral Antidepressant in Elderly Subjects With Treatment-resistant Depression
4. NCT02497287 - ESKETINTRD3004 - An Open-label, Long-term, Safety and Efficacy Study of Intranasal Esketamine in Treatment-resistant Depression
5. NCT02493868 - ESKETINTRD3003 - A Randomized, Double-blind, Multicenter, Active-Controlled Study of Intranasal Esketamine Plus an Oral Antidepressant for Relapse Prevention in Treatment-resistant Depression
6. NCT01998958 - ESKETINTRD2003 - A Double-Blind, Doubly-Randomized, Placebo-Controlled Study of Intranasal Esketamine in an Adaptive Treatment Protocol to Assess Safety and Efficacy in Treatment-Resistant Depression (SYNAPSE)
7. NCT02133001 - ESKETINSUI2001 - A Double-blind, Randomized, Placebo Controlled Study to Evaluate the Efficacy and Safety of Intranasal Esketamine for the Rapid Reduction of the Symptoms of Major Depressive Disorder, Including Suicidal Ideation, in Subjects Who Are Assessed to be at Imminent Risk for Suicide
8. NCT03039192 - 54135419SUI3001 - A Double-blind, Randomized, Placebo-controlled Study to Evaluate the Efficacy and Safety of Intranasal Esketamine in Addition to Comprehensive Standard of Care for the Rapid Reduction of the Symptoms of Major Depressive Disorder, Including Suicidal Ideation, in Adult Subjects Assessed to be at Imminent Risk for Suicide
9. NCT03097133 - 54135419SUI3002 - A Double-blind, Randomized, Placebo-controlled Study to Evaluate the Efficacy and Safety of Intranasal Esketamine in Addition to Comprehensive Standard of Care for the Rapid Reduction of the Symptoms of Major Depressive Disorder, Including Suicidal Ideation, in Adult Subjects Assessed to be at Imminent Risk for Suicide
10. NCT02918318 - 54135419TRD2005 - A Randomized, Double-blind, Multicenter, Placebo-controlled Study to Evaluate the Efficacy, Safety and Tolerability of Fixed Doses of Intranasal Esketamine in Japanese Subjects With Treatment Resistant Depression
11. NCT01640080 - ESKETIVTRD2001 - A Double-Blind, Double-Randomization, Placebo-Controlled Study of the Efficacy of Intravenous Esketamine in Adult Subjects With Treatment-Resistant Depression
12. NCT03434041 - ESKETINTRD3006 - A Randomized, Double-blind, Multicenter Active-controlled Study to Evaluate the Efficacy, Pharmacokinetics, Safety and Tolerability of Flexible Doses of Intranasal Esketamine Plus an Oral Antidepressant in Adult Subjects With Treatment-resistant Depression

**What type of data are you looking for?:** Individual Participant-Level Data, which includes Full CSR and all supporting documentation

### Research Proposal

### Project Title

Sex Differences in the Antidepressant Effects of Ketamine

#### Narrative Summary:

In 2019, after several successful clinical trials, ketamine was approved to treat these patients that do not respond to treatments by the FDA. Men and women may have different response rates to ketamine, as biological sex and gender are critical factors in diagnosing depressive disorders. Indeed, more women are diagnosed with MDD than men and present with greater suicidal ideation. However, it remains unclear if there are sex-dependent responses to ketamine. The present project aims to determine if there are differences in ketamine effectiveness and side effects by aggregating data from completed clinical studies (using a meta-analysis).

**Scientific Abstract:**

Background: Ketamine, an N-methyl-D-aspartate receptor (NMDAR) antagonist, is effective in patients with major depressive disorder (MDD) that are resistant to first-line therapies. Ketamine rapidly ameliorates depressive and suicidal symptoms. Preclinical literature suggests that there are sex differences in ketamine's effects, whereas this observation is inconsistent in clinical studies, although there is no meta-analysis.

The incidence of MDD is much greater in females than males. For example, in Canada, MDD is diagnosed in 5.8% of women and 3.6% of men. Furthermore, suicidality, a common feature of MDD, was also discovered to vary by sex. The risk of suicide attempts is higher for females, but the incidence of completed attempts is higher for males.

Given the variability in MDD symptomatology in males and females, as well as the disorder's greater frequency in females, it is vital to understand if ketamine's antidepressant and adverse side effects vary by sex.

Objective: To evaluate whether female and male MDD patients respond differently to ketamine treatments, we aim to perform a meta-analysis of clinical trial data to determine if effectiveness, suicidality and adverse effects differ between male and female patients.

Study Design: Meta-analysis of randomized clinical trials with data disaggregated by sex.

Participants: MDD patients 18 years and older. Biological sex needs to be reported to be included in the meta-analysis.

Primary and Secondary Outcome Measures: Primary outcome measures are the severity of depressive symptoms, suicidality scores and incidence of adverse effects.

Statistical analyses: Meta-analysis using STATA.

**Brief Project Background and Statement of Project Significance:**

Ketamine, an N-methyl-D-aspartate receptor (NMDAR) antagonist, has been found to relieve depressive and suicidal symptoms in patients with treatment-resistant depression (TRD), a classification of major depressive disorder (MDD). TRD has been associated with greater severity, duration, likelihood of comorbid conditions and risk of recurrence. (Culpepper, 2013) MDD occurred in 5.8% of women and 3.6% of men in Canada in 2012. (Albert, 2015) Furthermore, it has been observed that suicidality, a common feature of MDD, was discovered to vary by sex. The risk of suicide attempts is higher for females but the incidence of completed attempts in higher for males. (DSM–5; American Psychiatric Association, 2013)

A literature review of the preclinical literature (Ponton et al., 2022) suggests that there are sex differences in ketamine's antidepressant effects, with females responding more strongly to ketamine than males in 10 out of 23 studies. This observation is inconsistent in clinical studies. A recent systematic review of the clinical literature (Benitah et al., 2022) revealed that only 4 out of 27 studies presented sex differences in ketamine antidepressant response, with females responding somewhat more strongly. Given the variability in suicidality in male and female MDD patients, as well as the disorder's greater frequency in females, it is vital to understand if ketamine's antidepressant features may vary by sex in the clinical framework.

Ketamine rapidly reduces depressive symptoms and suicidal thoughts in MDD and TRD patients, either in its racemic form or as one of its two enantiomers, esketamine (S+) and arketamine (R-). (Abdallah et al., 2015; Tizabi et al., 2012; Saland et al., 2017; Niciu et al., 2019) Nevertheless, there remains more to understand about ketamine, such as its mechanism of action, its effects on select symptoms of depressive disorders and how these features interact with biological sex. Clinical trials with esketamine contributed to the YODA Project are thus excellent data sets to implement in a meta-analysis designed to better understand whether the effectiveness of ketamine on depression scores and suicidality, as well as adverse effects, differ between male and female patients.

**Specific Aims of the Project:**

The aim of this study is to determine if there are any sex differences in the antidepressant response to ketamine in clinical MDD samples, including suicidality and side effects. A meta-analysis and literature review is being performed to answer this question. Online scientific databases such as PubMed and PsycINFO were searched for studies using ketamine, with the strict requirement that results be split by sex for comparison purposes. Preclinical studies suggest that there is a greater sensitivity to ketamine in females than in males. We thus predict that female participants will have a greater response to ketamine in depressive symptom and suicidality alleviation than male participants.

**What is your Study Design?:**

Meta-analysis (analysis of multiple trials together)

**What is the purpose of the analysis being proposed? Please select all that apply.**

New research question to examine treatment effectiveness on secondary endpoints and/or within subgroup populations

New research question to examine treatment safety

Participant-level data meta-analysis

Meta-analysis using data from the YODA Project and other data sources

### Research Methods

**Data Source and Inclusion/Exclusion Criteria to be used to define the patient sample for your study:**

Inclusion criteria are that ketamine needed to be administered strictly to MDD patients, data needed to be reported by sex, patients needed to be 18 years of age or older, and all studies needed to be randomized control trials.

**Primary and Secondary Outcome Measure(s) and how they will be categorized/defined for your study:**

There are three primary outcome measure for this study. The first is the severity of depressive symptoms following treatment, as assessed by questionnaires such as the Montgomery–Åsberg Depression Rating Scale (MADRS) and the Quick Inventory of Depressive Symptomatology – Self-Report (QIDS-SR). The second primary outcome is suicidality scores (e.g., suicidal ideation, suicide attempts) following treatment, assessed by questionnaires like Beck's Scale for Suicidal Ideation (SSI) and the Clinical Global Impression of Severity of Suicidality Revised version (CGI-SS-r). The third primary outcome is the incidence of adverse effects (e.g., drowsiness, psychotomimetic symptoms such as hallucinations), following treatment, also assessed by questionnaires such as the Brief Psychiatric Rating Scale (BPRS) and the Young Mania Rating Scale (YMRS).

**Main Predictor/Independent Variable and how it will be categorized/defined for your study:**

The main independent variable for this study is participant biological sex. We will use self-report information from the original studies to classify participants by sex.

**Statistical Analysis Plan:**

All aim-relevant data extracted from studies will be graphed in STATA using a forest plot, which is to be disaggregated by sex. Funnel plots and the trim and fill method will then be generated to determine the severity of publication bias across all included studies. Afterwards, study influence will be assessed using the metainf command to discover which studies may be responsible for skewed meta-analysis outcomes. A new sex-disaggregated forest plot will then be generated excluding studies with high publication bias. All procedures listed will be performed three times for each of the three primary outcomes.
Software Used:
STATA

**Project Timeline:**

Project start date: September 1, 2022; meta-analysis already in progress and at the data extraction stage. Eligible studies did not all disaggregate findings by sex and authors were contacted to request access to the data. One of these authors referred us to the YODA Project for requesting data from additional studies. The data requested from the YODA Project is to be included in the present meta-analysis.

Anticipated analysis completion date: April 15, 2023

Anticipated date manuscript drafted: May 31, 2023

Anticipated first submission for publication: July 1, 2023

Anticipated date for reporting results back to the YODA Project: September 1, 2023

**Dissemination Plan:**

We plan to write a manuscript with the meta-analysis results. This manuscript will be reposited in bioRxiv, then submitted for publication to an open access biomedical journal.

.misc-fixes { display: none; } #admin-region { z-index: 9999999; } #admin-menu { z-index: 99999999; }
